# Sensitive detection of copy number alterations in samples with low circulating tumor DNA fraction

**DOI:** 10.1101/2024.05.04.24306860

**Authors:** Markus Mayrhofer, Rebecka Bergström, Venkatesh Chellappa, Anastassija Kotsalaynen, Sarath Murugan, Alessio Crippa, Bram De Laere, Karen Urtishak, Karina Dalsgaard Sorensen, Kavita Garg, Usha Singh, Martin Eklund, Henrik Grönberg, Johan Lindberg

## Abstract

Copy number analysis is an important aspect of cancer genomics that enables identification of activated oncogenes, inactivated tumor suppressor genes and genome-wide signatures such as homologous recombination deficiency and the tandem duplication phenotype. Despite continuous development of copy number algorithms, the current sensitivity to detect clinically relevant focal alterations is poor if the cancer DNA fraction is low. This is particularly challenging for analysis of circulating tumor DNA (ctDNA) as it is not possible to know the cancer DNA fraction in advance or, as for tissue, macrodissect to increase the cancer DNA fraction. Here, we combine a novel algorithm (Jumble) with a tailored gene panel design and selected reference samples that achieve sensitive and highly specific detection of clinically relevant copy number alterations with limits of detection at 1-2% ctDNA fraction for amplifications and 4-8% for homozygous deletions. Jumble lowers the ctDNA fraction required for detection of homozygous deletions 3-6 times compared to commercial alternatives. Jumble is freely available as an R script and container, ready for integration into bioinformatic pipelines.

## Background

Analysis of tumor DNA is important in cancer research and clinical management^1,2^. Tumor DNA can be obtained from biopsy or surgical resection of tumor tissue and from blood plasma as circulating tumor DNA (ctDNA). An important feature of DNA extracted from tumor tissue is that the original sample typically contains a significant fraction of non-cancerous cells which will not carry the somatic alterations that drive the disease. Similarly, ctDNA comprises a fraction of the total cell-free DNA (cfDNA), which originates to a large extent from white blood cells^3^. As the cancer DNA fraction required for detection of different types of somatic alterations varies, sequencing analysis may result in inconclusive biomarker results if purity is low^4,5^. High sequencing depth can attain high sensitivity despite a low cancer DNA fraction, but is not affordable for whole genome sequencing. Therefore, especially for solid cancers, targeted sequencing approaches dominate biomarker analysis, both in the academic and routine diagnostic setting. Targeted or so called panel-based approaches have converged at applying assays with an uneven distribution of targets throughout the genome to enable detection of different types of somatic alterations such as gene fusions, copy number alterations (CNAs) and small mutations^6^.

For copy number analysis, the sequencing depth, (i.e. a number of DNA reads or fragments) at predefined intervals (bins) is used to estimate the DNA abundance in a sample, throughout a reference genome, and requires correction for variables affecting depth such as GC content bias. This can be achieved using regression models for known sources of bias. A patient-matched normal sample or a reference pool of non-aberrant samples can be used to remove additional systematic variation. This approach, initially developed for microarray data, has evolved to apply complex representations of the reference pool using for example principal component analysis^7–10^. The resulting corrected sequencing depth is a DNA abundance ratio centered at 1 or a log ratio centered at 0.

Despite the plethora of bioinformatic software tools and sequencing approaches, sensitive detection of certain copy number alterations is currently an unmet challenge, exemplified by the recent approval of PARP inhibitors in metastatic castrate resistant prostate cancer (mCRPC) for patients with inactivating mutations in *BRCA1* and *BRCA2*^*11,12*^. Cancers with somatic homozygous deletion of *BRCA2* demonstrate superior responses compared to other types of alterations^13–15^. Ironically, detection of homozygous deletions require the highest purity of all somatic alteration types. Both commercial assays and academic initiatives have reported a homozygous deletion limit of detection threshold of approximately 0.2 to 0.5 ctDNA fraction ^4,5,16–18^. As the majority of men with first line mCRPC have a ctDNA fraction below 0.2, the biomarker status for *BRCA1* and *BRCA2* will remain inconclusive in 50 - 75% of mCRPC patients depending on assay performance^5^.

The Prostate Biomarkers (ProBio) trial^19^ is an outcome-adaptive biomarker driven platform trial in men with metastatic hormone sensitive prostate cancer (mHSPC) and mCRPC. In ProBio, a predefined set of somatic and germline alterations are categorized into biomarker signatures which are evaluated in multiple experimental arms versus standard of care, defined as physician’s choice of treatment. Biomarker signatures are assessed through synchronous targeted sequencing of ctDNA and germline DNA. For ProBio, we have developed a prostate-specific panel design (ProBio panel) which allows for cost efficient targeted interrogation of the cancer genome at the expected range of ctDNA fractions. However, sensitive identification of treatment relevant homozygous deletions in genes such as *BRCA2* remains challenging. Therefore we have developed Jumble, a novel computational method that in combination with the panel design achieves high sensitivity to detect homozygous deletions and other types of CNAs at low ctDNA fractions.

## Methods

Extracted germline DNA (gDNA) and/or plasma cfDNA was obtained from three studies, all analyzed with the ProBio panel. **Dataset 1**: cfDNA and matched gDNA from the ProBio trial (ClinicalTrials.gov identifier: NCT03903835, www.probiotrial.org). **Dataset 2**: cfDNA from the MAGNITUDE trial (ClinicalTrials.gov identifier: NCT03748641) that were part of a blinded evaluation of the Resolution ctDx HRD assay from Resolution Biosciences with the academically established ProBio assay.^5^ All homozygous deletions in BRCA2 were detected by both assays (separate manuscript in preparation). Upon request Resolution Biosciences kindly provided homozygous PTEN calls which were all re-identified by the ProBio assay. **Dataset 3:** A Danish retrospective cohort, described in Nørgaard et al^20^ for which low-pass whole genome sequencing data was available on the same plasma samples analyzed with the ProBio panel.

All relevant ethical guidelines have been followed, all necessary IRB and/or ethics committee approvals have been obtained, all necessary patient/participant consent has been obtained and the appropriate institutional forms archived. Dataset 1 (ProBio, ClinicalTrials.gov identifier: NCT03903835) and dataset 2 (Magnitude, ClinicalTrials.gov identifier: NCT03748641) originate from international randomized clinical trials. For both studies local IEC/IRB approval was obtained before initiating the study. E.g. in Sweden the studies were approved by the Swedish Ethical Review Authority. Dataset 3, the study was approved in Denmark by the National Committee on Health Research Ethics. Dataset 1-3: all patients provided written informed consent.

Library preparation was performed with Kapa DNA HyperPrep (Roche) using UMI xGen CS Adapters (IDT). In-solution hybridization based capture was applied to enable targeted sequencing using baits (120 bp oligos) obtained from Twist Bioscience. The baits, defining the ProBio panel (v3), target regions in the human genome to enable comprehensive cost efficient analysis of the prostate cancer genome. Briefly, the ProBio panel enables detection of point mutations (78 genes), structural variants by intronic sequencing (11 genes), increased bait density to improve copy number alteration sensitivity (20 genes), microsatellite instability (MSI) and hypermutation (Table S1). The design targeted approximately 3000 common SNPs which enables genome-wide copy number alteration profiling and ploidy assessment. Illumina paired-end sequencing (2 × 150 bp) was performed on the NovaSeq system (Illumina). For tumor and germline DNA, ≥80 × 10e6 and ≥15 × 10^6^, respectively, read pairs were ordered (Table S2).

Sequencing data underwent processing using the in-house bioinformatics pipeline AutoSeq, which integrates several widely-used tools and also in-house developed tools. The pipeline begins with raw reads in FASTQ format. These reads are trimmed using Skewer^21^ to eliminate adapters. Duplex unique molecular identifiers (UMIs) are then extracted from the trimmed FASTQ files and annotated to the raw reads in an unmapped BAM format using fgbio’s FastqToBam (http://fulcrumgenomics.github.io/fgbio/). The Picard tool SamToFastq (http://broadinstitute.github.io/picard), converts these unmapped BAM files back to FASTQ, which serves as input for BWA. Subsequent alignment is performed using BWA MEM^22^. The mapped raw BAM is then annotated with UMI tags from the unmapped BAM using Picard’s MergeBamAlignment. GATK3’s RealignerTargetCreator and IndelRealigner^23^ are then used to realign the aligned raw reads around indels, producing the final raw BAM. To construct consensus reads, raw reads are grouped by mapping position and UMI using fgbio’s GroupReadsByUmi. Consensus reads are then generated from these groups with fgbio’s CallDuplexConsensusReads. The resulting consensus reads, packed with detailed BAM tags, undergo a similar alignment process as the raw reads. This includes conversion from unmapped BAM to FASTQ using Picard’s SamToFastq, alignment via BWA MEM, and annotation with consensus read information from the unmapped BAM using Picard’s MergeBamAlignment. The aligned consensus reads are realigned around indels using GATK3. Finally, the consensus reads are refined using fgbio’s FilterConsensusReads and ClipBam to remove any overlapping read pairs.

Further steps in the AutoSeq pipeline include purity and ploidy assessment and variant calling and annotation followed by manual curation of data^5^. Calling of small somatic variants, i.e. single nucleotide variants (SNVs) and insertions/deletions (indels), is performed with two variant callers: GATK Mutect2^23,24^ and HMF Tools SAGE^25^ for tumor sample and matching germline sample. Results are merged with SomaticSeq^26^ and annotated with Ensembl Variant Effect Predictor (VEP)^27^. Variants called by ≥1 caller and with impact high or moderate are included for manual curation. Calling of small germline variants is performed with GATK HaplotypeCaller^23,28^, and results are annotated with VEP. Filters applied before considering variants for manual curation of potentially pathogenic germline variants include: 1) Impact high or moderate 2) If consequence is missense: must have a ClinVar^29^ annotation pathogenic or likely pathogenic.

Structural variants are called with GRIDSS^30,31^ and in-house algorithm SVcaller^5^. Filtering is performed before considering variants for manual curation keeping variants with: 1) ≥3 supporting reads; 2) overlap with any exon; 3) overlap of either breakpoint with a target in the panel (e.g. TMPRSS2-ERG introns for fusion detection). Purity and ploidy analysis is performed using PureCN^9^, which takes CNA and small variant data as input. Where that result is deemed incorrect based on manual inspection of CNAs and SNP allele ratio, somatic mutation VAFs are used for purity estimation while ploidy is estimated based on the copy number profile.

Interpretation and visualization of variants is undertaken in an in-house developed software, Curator, together with the integrated genomics viewer (IGV)^32^. For cases without any available germline DNA, healthy donor DNA is applied as a germline DNA reference. To enable identification of somatic variants in cases without matching germline DNA, all small variants with >0.01 population frequency in the Genome Aggregation Database (gnomAD)^33^ are assumed to be germline variants. The remaining variants are manually investigated. Each variant of apparent relevance (mutations, copy-number alterations, and structural variants) is inspected to identify artifacts^34^, cancer driver- or passenger alterations and to confirm the cancer DNA fraction estimate.

## Results

Here we present a copy number analysis algorithm, Jumble, and its performance with a targeted hybrid-capture gene panel (the ProBio panel), developed for the ProBio trial^35^. We sequenced cell-free DNA from three different cohorts of metastatic prostate cancer with the ProBio panel; Dataset 1 from the ProBio trial^19,36^, N=193 including patient-matched samples from different timepoints; Dataset 2 from the MAGNITUDE trial^37^, N=266 including samples with known *BRCA2* and *PTEN* homozygous deletions; and Dataset 3 (Danish retrospective mCRPC cohort), with matching shallow whole genome sequencing data (N=90 with another N=134 with WGS only). A reference sample set for Jumble was created using cell-free and whole-blood DNA from healthy donors and Dataset 1 samples with no evidence of cancer DNA (i.e. cancer-related mutations, N=78).

Jumble is available as an executable R-script with a Docker/Singularity container (https://github.com/ClinSeq/jumble). It applies PCA to identify features associated with systematic variation, i.e. noise, in reference samples, and corrects for variability associated with these features to remove noise from query (to be analyzed) samples (**Supplementary Figure 1**). While designed to address challenges with the ProBio panel design (and similar panels), Jumble is suitable for any human sequencing data including panels of any size, exomes and whole genome sequencing.

The ProBio panel design, together with Jumble, attains high sensitivity and specificity for copy number alterations such as homozygous deletions (deletions of both copies of a gene, **Supplementary Figure 2**), of tumor suppressor genes such as *BRCA2* and *PTEN*, and amplifications of oncogenes such as *AR*. Common SNPs are targeted genome-wide about 1 Mb apart. Clinically relevant genes are interrogated through sparse (exons only) or dense (full gene body) targeting. Selected genes, commonly altered via copy number alterations, are supplemented with additional targets that serve as control regions located far enough from the gene not to be typically included in a homozygous deletion^38^. An overview of the ProBio panel is presented in **Figure 1** with additional details in **Supplementary Table 1**.

**Figure 1.**
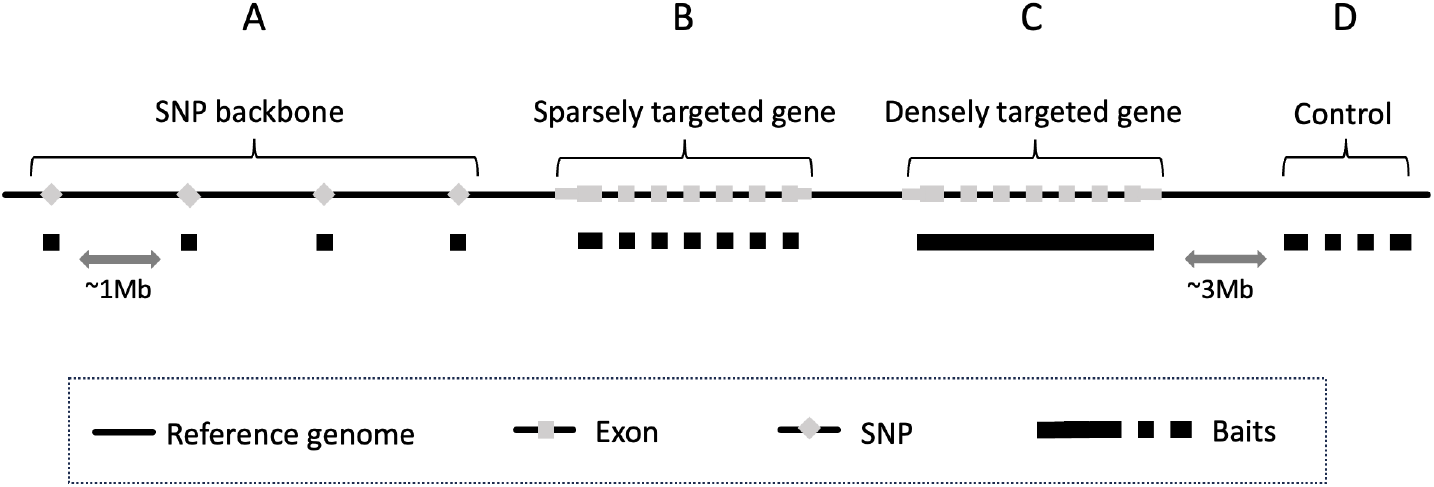
General target or oligo capture location strategy. **A)** SNPs are targeted (captured using baits; 120 bp oligos) throughout the reference genome, separated by about 1 million bases to enable genome wide copy number alteration analysis. **B)** Cancer-relevant genes are sparsely targeted with baits at all or selected exons only. **C)** Some genes where copy number alterations and structural variants are of particular importance are densely targeted with most or all of the gene body tiled with adjacent baits. **D)** For selected genes, additional control regions upstream and downstream of the gene, are targeted to increase focal copy number breakpoint resolution.

A challenge with the ProBio panel is that densely targeted gene bodies correspond to a large fraction (0.53) of the total target footprint. These gene bodies may differ systematically from other targeted genomic regions in features such as GC content with many gene bodies typically GC poor relative to intergenic sequence^39^.In addition, there may be systematic differences between the sequence coverage of densely and sparsely targeted genomic regions. When such densely targeted genes harbor deep deletions or high amplifications, a dependency may emerge between the systematic variability in sequence coverage and the signal representing a copy number alteration. Conventional correction, e.g. for GC bias, would remove signal in the affected genes, by mistaking signal for bias, and introduce errors in other genes. Jumble is designed to counter this issue while efficiently and effectively identifying and correcting for systematic noise (**Figure 2**).

**Figure 2.**
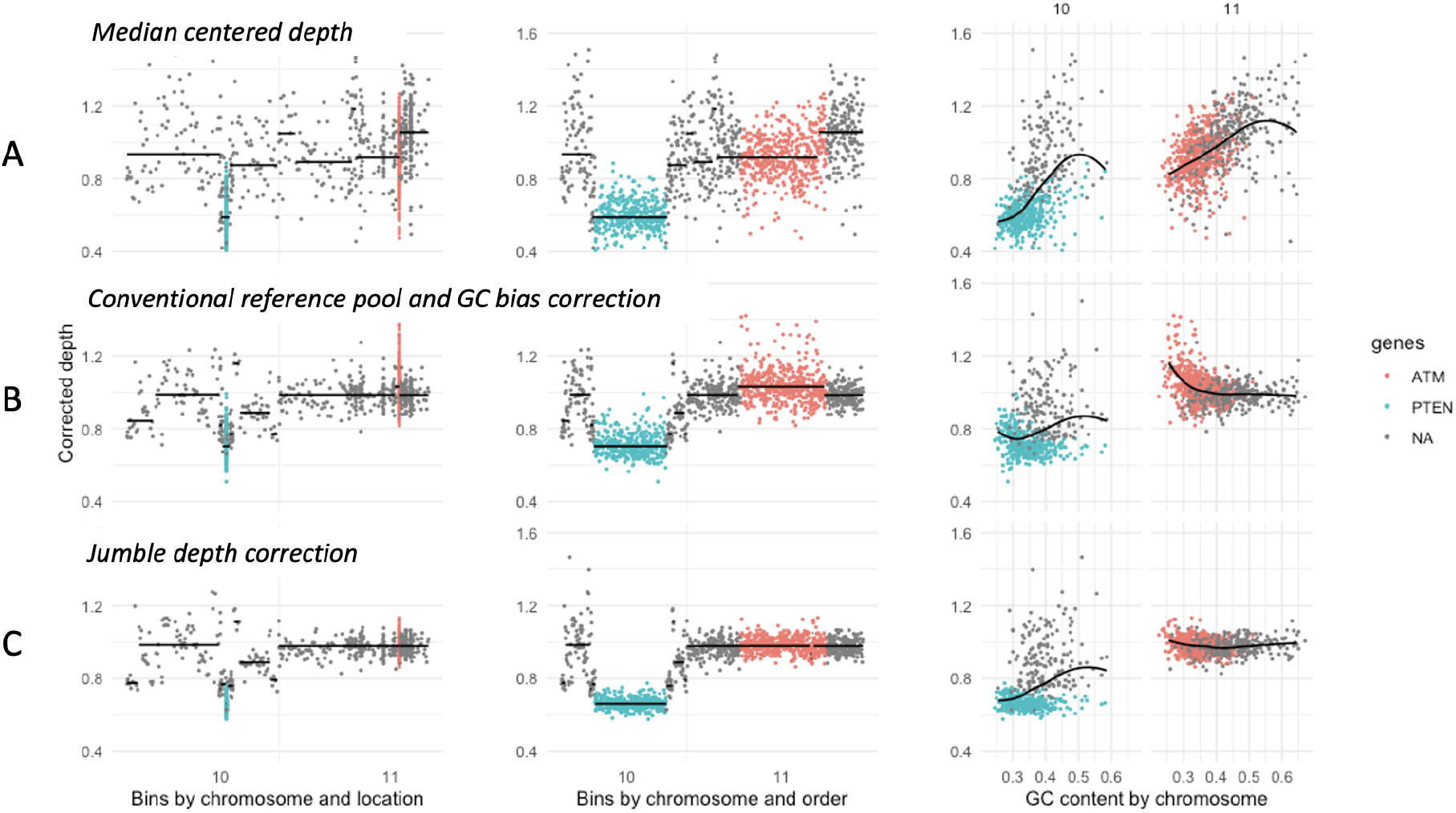
Sequencing depth correction overview. Chromosomes 10-11 are shown for a prostate cancer cell-free DNA sample presenting with somatic homozygous deletion of PTEN and no copy number alteration of ATM. The gene bodies are densely targeted for these genes. **A)** Median centered, otherwise uncorrected sequencing read depth, per genomic bin or location, is a crude estimate of DNA abundance in the sample analyzed. **B)** Conventional depth correction includes comparing the depth of each bin to a set (or a “best-fit” representation) of one or more reference samples, as well as correcting sequencing depth for any dependency with sequence features associated with sequencing depth bias, here exemplified with GC content. This can also introduce error in the result when there is a dependency between signal (here represented by the deletion of PTEN) and a feature used for correction (here represented by the low GC content of PTEN). Correction without mitigating the dependency between signal and noise distorts the resulting corrected depth, represented by an erroneous increase in corrected depth over PTEN and ATM bins with low-GC content. **C)** The Jumble algorithm removes noise while reducing the potential for introducing such errors, enabling copy number analysis to proceed with improved sensitivity and specificity.

The Jumble copy number analysis algorithm features correction of sequencing depth in query samples subjected to the analysis based on sequencing depth variation in a set of non-aberrant reference samples, as described in detail in **Supplementary Methods**. Briefly, PCA is applied to the reference set to compute latent features representing orthogonal observations of sequencing depth variability in the reference set, with a score for each bin and feature. Query sample sequencing depth is subsequently corrected using robust regression models of sequencing depth from feature scores. Importantly, the potential for densely targeted genes to perturb a model as shown in **Figure 2** is regulated by restricting regression model training to a subset of data points, called the *training* subset, effectively down-weighting some or all individual genes to cap their influence on the model. Segmentation of corrected depth is performed using circular binary segmentation^40^. Segments are annotated with known cancer genes and for each sample a visual summary is also generated (**Supplementary Figure 3**).

A noise estimate based on median absolute pairwise difference indicated that Jumble consistently outperforms conventional noise correction (Reference pool median subtraction and LOESS-based GC content correction, **Supplementary Figure 4**). To investigate whether copy number profiles and particularly focal deletions were detected reliably in samples with low ctDNA fraction, we investigated samples in Dataset 1 where ctDNA was analyzed from the same patient at different time points and where at least one featured a high ctDNA fraction (≥0.2). Manual curation and review of both corrected depth and SNP allele ratio profiles were applied. Copy number profiles, where observable, were consistently similar between patient-matched samples. In all samples, including low-fraction samples, where a homozygous deletion was indicated and there was a patient-matched sample available with higher ctDNA fraction, the indication was confirmed. Examples are shown in **Figure 3**. Residual noise and waviness in the corrected depth were generally less prominent at lower ctDNA fraction.

**Figure 3.**
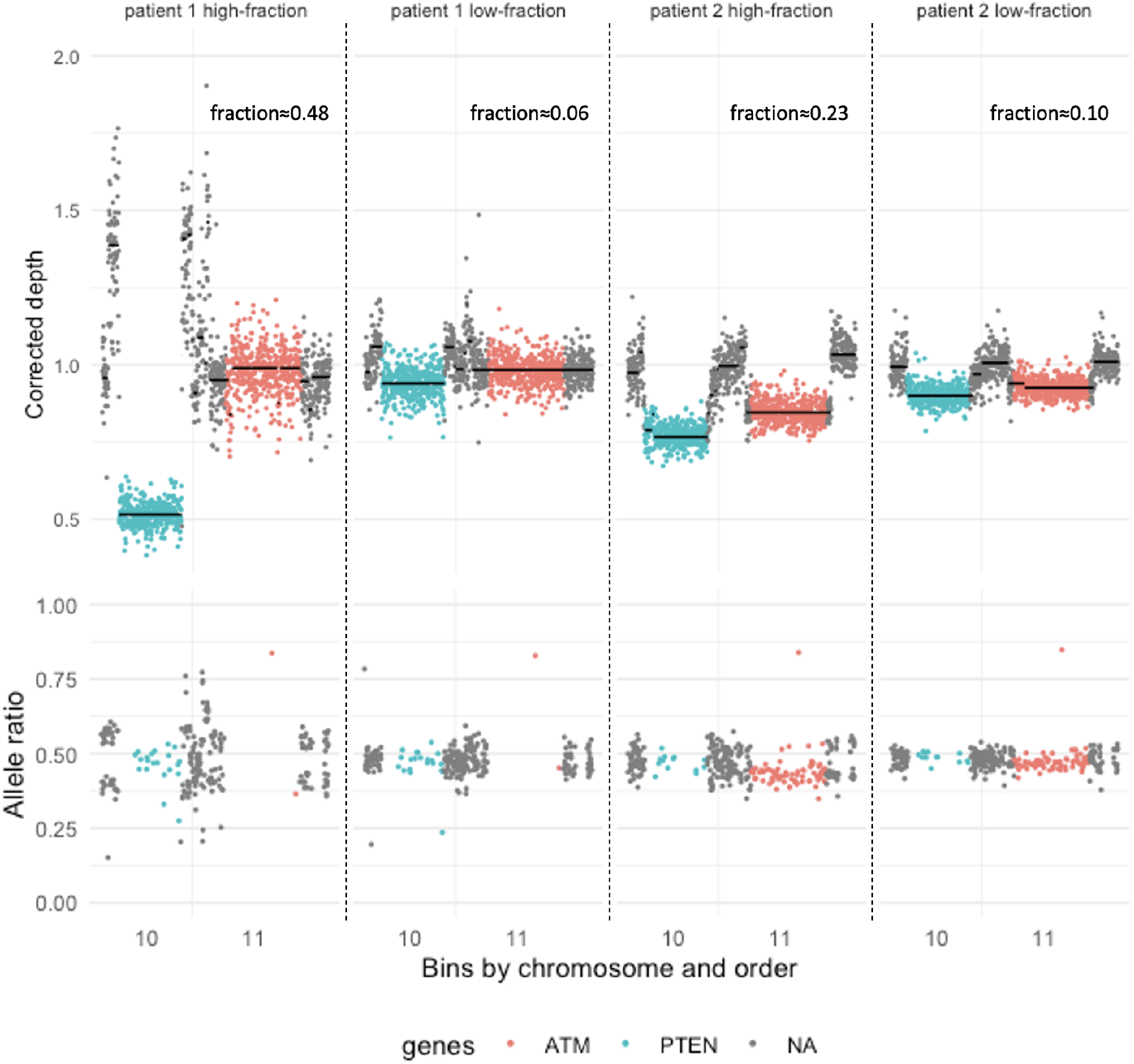
Matched cfDNA profiles at different time points. Upper panel: Corrected depth by order of chromosomal location for targeted chromosome 10-11 bins, for two sets of patient-matched ctDNA samples. **Lower panel:** Corresponding heterozygous SNP allele ratio. Each sample features somatic homozygous deletion of the PTEN gene, here indicated by the low corrected depth and balanced (about 0.5) SNP allele ratio. ATM is not homozygously deleted in either patient but hemizygously deleted in Patient 2. In sample 1, ATM does not feature heterozygous SNPs. In Patient 1 and with higher ctDNA fraction, waviness is prominent in the corrected depth, but this is much less of an issue at the lower ctDNA fraction, where systematic variability is better matched by the ctDNA-negative reference pool. All observations in the dataset of low ctDNA fraction (ca 0.05-0.20) and a clinically relevant homozygous deletion, and where a high-ctDNA fraction sample was available (≥0.20) from the same patient, were indicated as true deletions.

The performance of assays for analysis of ctDNA is commonly determined by the use of commercially available reference samples harboring e.g. mutations which was also done for the ProBio assay (**Supplementary figure 5-6**). Such samples do not exist for CNAs. Instead, cfDNA samples (Dataset 2) with known homozygous deletions (BRCA2, N=12; PTEN, N=24), identified by an orthogonal assay, were applied together with other genes (e.g. CHD1, N=8) for which de novo homozygous deletion detection was performed with Jumble. Samples in Dataset 2 with no evidence of cancer-related mutations were considered equivalent to ctDNA-negative or normal cfDNA, and used to create an *in silico* dilution series encompassing ctDNA fractions 0-0.20 and median sequence depths of 1000-4000X (sampling the normal DNA from more than one normal was necessary, described in detail in **Supplementary Methods**). Jumble was run on the diluted samples using the reference set generated from Dataset 1 (thus the reference set did not include any the ctDNA-negative samples used in the dilution series). An automated approach was used for the copy number calling procedure downstream of segmentation, without applying manual data curation. Briefly, a homozygous deletion was called if the gene, or a part of the gene, was segmented separately and with the segment(s) featuring a drop in corrected sequencing approximately similar to the theoretical expectation, given the DNA fraction and a homozygous deletion. Sensitivity estimates were computed, for a given ctDNA fraction and gene, using all diluted samples at that ctDNA fraction and with homozygous deletion of that gene. Specificity estimates were computed using all samples in the dilution series known not to harbor a homozygous deletion in the gene of interest, diluted to that ctDNA fraction. For *BRCA2* and *PTEN*, sensitivity reached about 50% at a DNA fraction of 0.04, and 90% at fractions of 0.08-0.12 and sequence coverage of 2000-3000 (**Figure 4)**. For *CHD1*, sensitivity was lower, which was expected as *CHD1* is not densely targeted in the ProBio panel. Specificity remained near 1 throughout the series, indicating that false positive calls can be avoided even at very low ctDNA fraction. Similar observations were made for the other included genes; these may be less reliable as only one or two samples per gene were available for the dilution series. (**Supplementary Figure 7)**.

**Figure 4.**
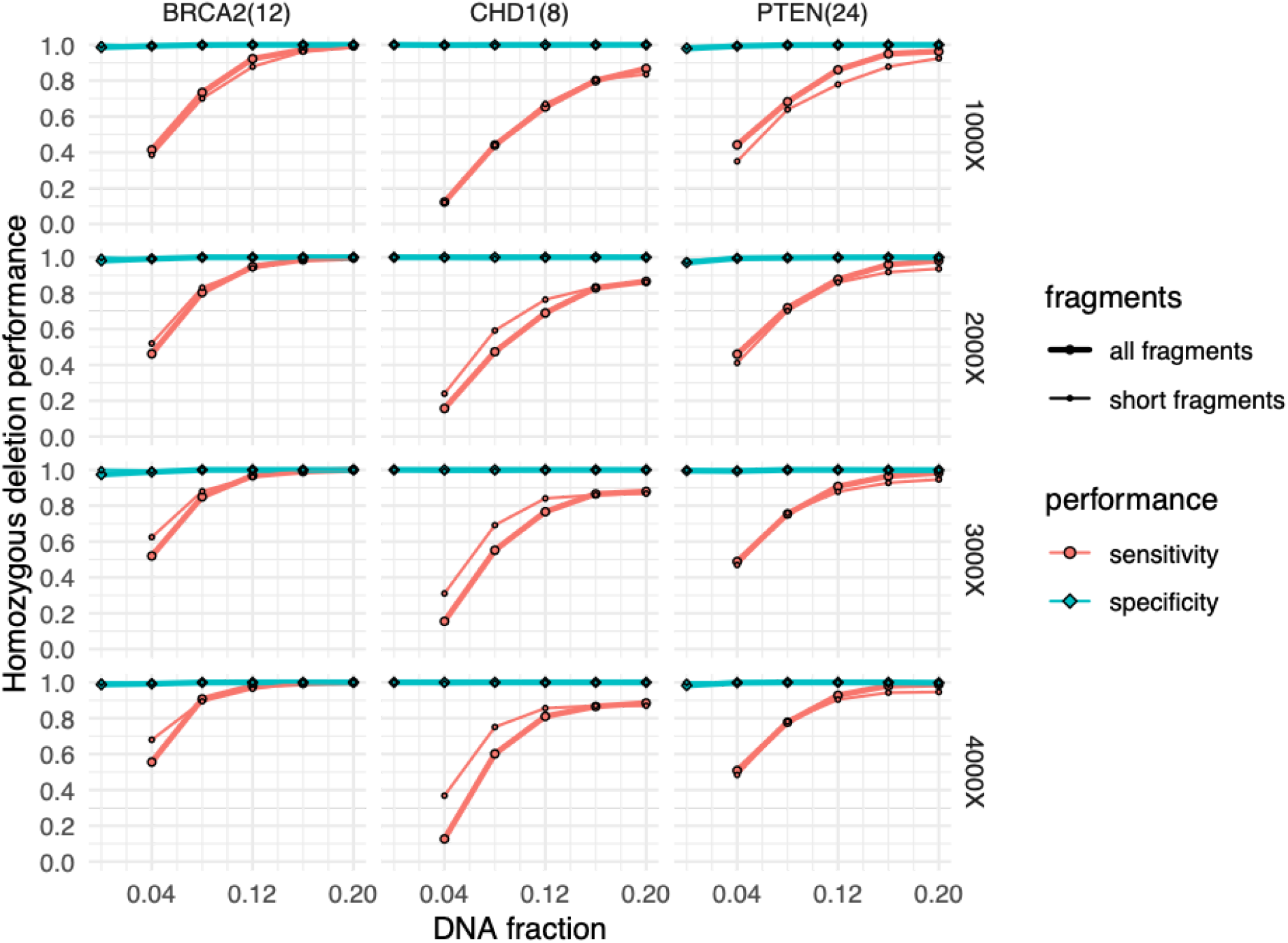
Sensitivity and specificity to detect homozygous deletions. An in silico dilution series of samples with known (PTEN, BRCA2) or high-confidence observation of homozygous deletion (CHD1), in 1-3 ctDNA-negative samples (as required to avoid oversampling) to sequencing depths of 1000-4000. Sensitivity and specificity estimates are shown for the included genes, by ctDNA fraction, fragment length and sequencing depth. Filtering sequence reads by fragment length (<150 bp) does largely not improve performance. The number of ctDNA samples with homozygous deletion available (=included) for generating dilutions in the experiment, by gene, is shown in parenthesis.

As ctDNA constitutes short cfDNA fragments, studies have shown that the apparent ctDNA fraction can be increased by filtering out longer sequence fragments from the analysis^41,42^. We performed the dilution series analysis separately using a fragment length filter (retaining fragment sizes <150 bases in both query and reference samples) to assess to what extent such filtering would affect our sensitivity and specificity estimates (**Figure 4 and 5, Supplementary Figure 7**). As expected, we observed an increase in the apparent ctDNA fraction, but the increase differed between genes, and more noise was observed due to lower remaining sequencing depth. Performance for homozygous deletion detection was generally similar, with only small differences observed throughout the range of genes, DNA fractions and sequencing depths, indicating that despite the apparent increase in ctDNA fraction as an effect of a fragment length filter, the net effect of removing longer fragment reads is not generally an improved performance for detecting focal deletions. An example of the result where focal deletion of *BRCA2* was successfully segmented at a ctDNA fraction of 0.04 and higher is shown in **Figure 5**.

**Figure 5.**
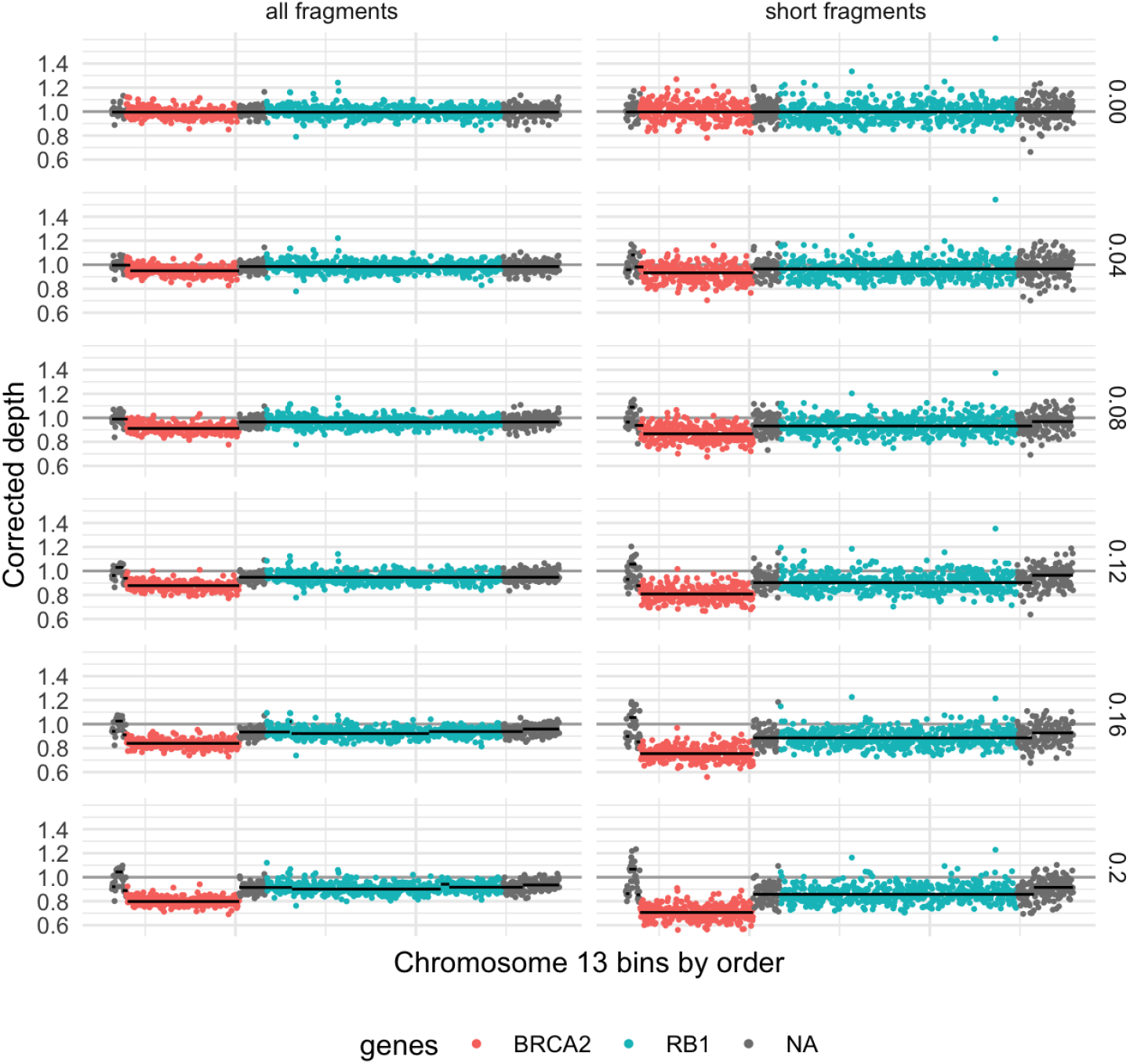
Homozygous deletions at different ctDNA fractions. Example of corrected depth and segments through chromosome 13 in a diluted sample (depth 2000X, ctDNA fraction 0-0.20) with BRCA2 homozygous deletion, and with nonfocal hemizygous deletion of RB1 and most of chromosome 13. The effect of analyzing the sample using only short DNA fragments is shown for comparison, leading to a combination of more noise and stronger signal.

Performance estimates for gene amplifications are challenging to generalize, as they range from focal to chromosome arm level and from a few extra copies to hundreds. The ProBio panel features dense targeting of the androgen receptor (*AR*) and the *AR* enhancer^43^. Both are commonly amplified in advanced prostate cancer due to evolutionary pressure from androgen deprivation therapy, with pronounced heterogeneity between subclones further complicating systematic assessment of sensitivity and specificity^44,45^.

Distributions of *AR* corrected depth were similar across Datasets 1, 2 and 3, as well as low-pass whole genome sequenced samples of Dataset 3, ranging from a modest decrease to about 30-fold increase relative to median of the non-AR chromosome X bins (**Supplementary Figure 8A)**. Healthy donor samples showed very small deviations in corrected depth. Matched comparison of *AR* corrected depth in Dataset 3 for the Probio panel and Jumble, against the same samples analyzed with shallow WGS and ichorCNA^46^, did not indicate any saturation effect for targeted sequencing and aligned well for commonly altered chromosome arms (8p and 8q) and for the AR gene and enhancer (**Supplementary Figure 8B)**.

Absolute copy number estimates of amplified genes require ploidy assessment which is error-prone for low-purity samples. We calculated ploidy-agnostic, ctDNA fraction-adjusted *AR* and *AR* enhancer copy number estimates in Dataset 1, assuming average ploidy of two. Co-amplification of the enhancer and the gene was typically observed, but with exceptions where *AR* and enhancer copy number estimates differed. This included amplifications of only the enhancer or gene, as previously reported in other studies^47^. The copy number estimates formed a continuous distribution ranging as high as approximately 30 copies, without indicating a separation between high- and low-level copy number alterations (**Figure 6**). Some apparent decreases in corrected depth were also observed, equivalent to putative copy loss events, but none of them indicated deletion of a whole copy (clonal deletion of one copy) and they were considered to be the effect of residual waviness. This apparent waviness was mostly prominent in samples with higher ctDNA fraction and it was not observed in samples from healthy donors (**Supplementary Figures 8, 9**).

**Figure 6.**
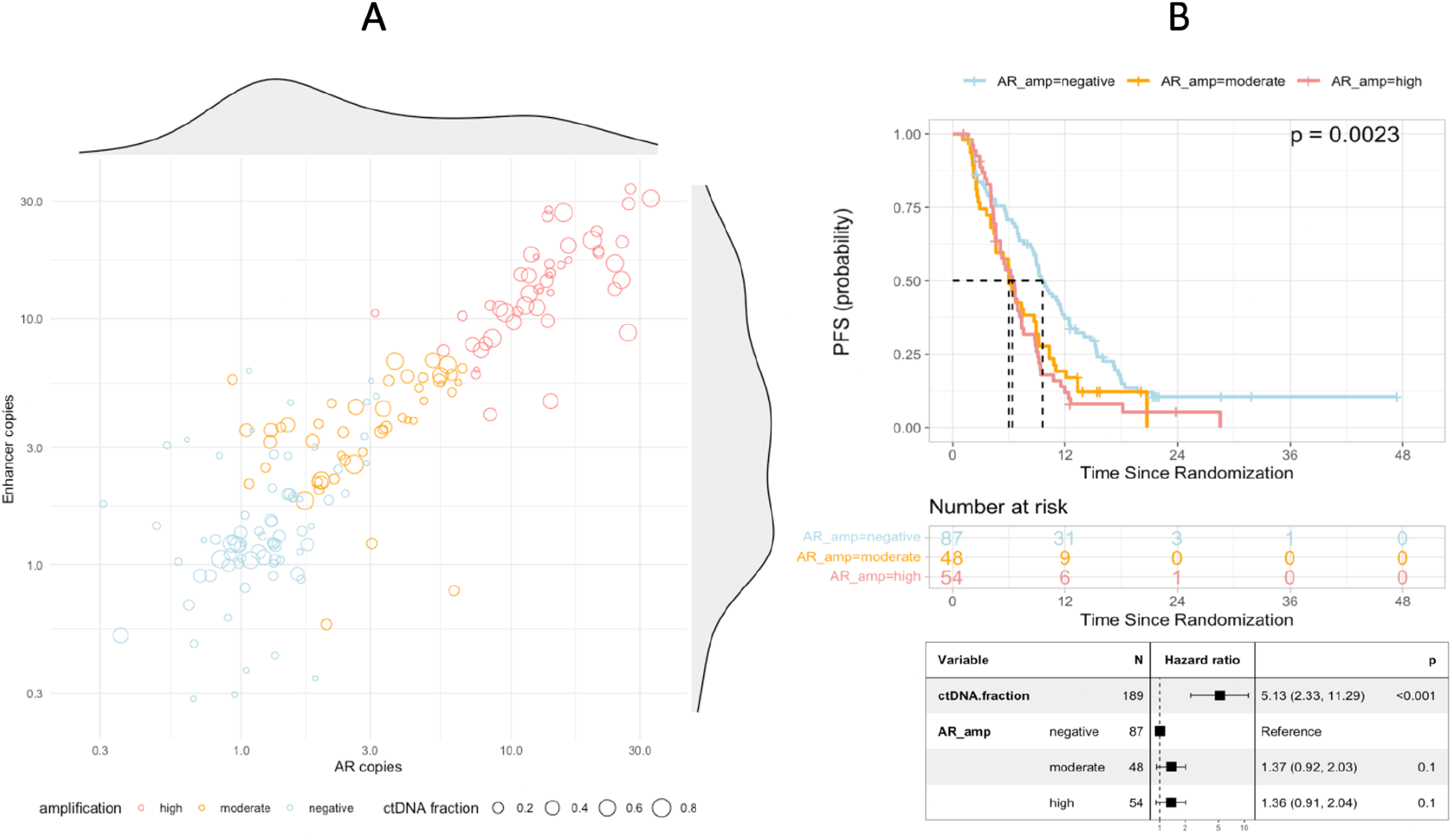
Detection of AR and AR enhancer amplifications. **A)** Ploidy-agnostic estimates of copy number, based on corrected sequencing depth, the ctDNA fraction, and relative to a ploidy of 2, of AR and the AR enhancer in Dataset 1, with density distributions. These copy number estimates can sometimes appear to be below 1 without indicating deletion, as the actual ploidy or average copy number can be above 2. Samples where AR or its enhancer are considered amplified, according to our criteria, are labeled as moderate (<7 copies) or high (≥7 copies) amplification. **B)** AR and/or AR enhancer amplification status in plasma cell-free DNA and progression-free survival. Samples with >1% ctDNA fraction (n=189) were selected from the ProBio trial (Dataset 1). Blood samples had been collected and subjected to isolation and analysis of circulating cell-free DNA (cfDNA) prior to starting a new line of systemic therapy, encompassing AR pathway inhibitors (n=73), taxane-based chemotherapy (n=110), or other systemic therapy (n=6). Upper: Kaplan-Meier analysis of progression-free survival, stratified according to AR and/or enhancer amplification status (negative, moderate, and high-level amplification). p-value is calculated via log-rank test. Lower: Multivariable Cox regression analysis of progression-free survival using baseline ctDNA fraction and AR and/or enhance amplification status (bottom). p-value is calculated via Wald test.

To pragmatically call *AR* and enhancer amplifications with high sensitivity and specificity, we set thresholds above the apparent amplitude of waviness: One or both of *AR* and enhancer median corrected depth were required to either a) exceed the expected and observed chromosome X medians by 50% (>0.75 with an X chromosome median value of 0.5), or b) exceed the expected and observed chromosome X medians by 10% (>0.55 with an X chromosome median value of 0.5*) and* exceed a mean of two copies per cancer cell (assuming average-diploid genome). In average-diploid genomes with a copy number of one throughout most of the X chromosome, this results in an expected sensitivity to detect amplifications averaging ≥11 copies per cell at a tumor DNA fraction of 1%, ≥6 copies per cell at a DNA fraction of 2%, and ≥2 copies per cell at a DNA fraction of 10% (**Supplementary Figure 9**).

Available progression-free survival data from Dataset 1 was to address the clinical validity of this approach. Observed *AR* amplifications were divided into high-level and modest amplifications, using the median copy number of observed amplifications (7 copies) as a cutoff^48^. The prognostic effect of *AR* amplification was found to be almost identical for high-level and modest AR amplification, thereby verifying acceptable specificity and clinical validity (**Figure 6B**).

## Discussion

The ProBio panel and copy number analysis software Jumble, presented here, was implemented as part of the genomic profiling applied in the ongoing ProBio trial. Typically, when evaluating cancer genomics assays, commercial reference samples are applied to evaluate the performance of variant calling. A challenge with calibrating copy number alteration detection performance is the lack of samples with known variants. Here, cfDNA samples with known homozygous deletions, verified by an independent technology, were applied to evaluate the performance of the ProBio panel and Jumble, showing a sensitivity to detect homozygous deletions as low as 4% for the homozygous deletions and 1-2% for amplification, whilst maintaining high specificity.

Homozygous deletions are always small, often spanning just a part of a gene and sometimes single exons. The results presented here serve to support that the ProBio panel and Jumble can be applied to call focal, presumed homozygous deletions at low ctDNA fraction without an inflated risk of type 1 error, as long as there is no deviation in technical quality. While the results are reassuring, manual review and curation remain in the standard procedure for the ProBio trial and are applied for all alterations of apparent relevance. The application of a fragment length filter for copy number analysis did, although an apparent increase in ctDNA fraction was observed as expected, not result in increased performance for detecting the relevant deletions.

With the copy number neutral reference sample set featuring primarily ctDNA-negative cfDNA samples, it matches the non-tumor cfDNA well, apparently resulting in less residual noise and waviness the lower the ctDNA fraction. This indicates that if higher-fraction ctDNA samples (without copy number alterations) were to be added to the reference set, a less wavy profile should be possible throughout the range of ctDNA fractions encountered. Such samples are not readily available however, and at high ctDNA fraction waviness does typically not resemble copy number alteration, and if it does, heterozygous SNP allele ratio can be used to support or contradict indications that may be the result of waviness. It can be assumed though, that if the reference samples are not well matched to the normal DNA component of query samples, the waviness can be a significant problem at any tumor DNA fraction.

Heterozygous SNP allele ratio is helpful as orthogonal (not subject to similar potential sources of error) verification that a putative copy number alteration is not actually waviness. Unfortunately, as exemplified by *ATM* in Sample 1, Figure 3, genomic regions the size of a single gene can be naturally homozygous due to the typical length of genomic haplotypes. Heterozygous SNPs can therefore not generally be relied upon to be present in a focal alteration. The SNP allele ratio can also be used with corrected sequencing depth to infer genomewide purity and ploidy estimates by fitting the observed data to a range of expected ploidies and purities, but this is prone to error below certain cancer DNA fractions, potentially therefore serving to worsen the limit of detection for relevant focal alterations. Our datasets and dilution experiment show that homozygous deletions and focal amplifications can be detected with high sensitivity and specificity, without requiring genome-wide absolute copy number estimates. Similar performance should be possible using e.g. tissue samples and other gene paneles, but the limit of detection would need to be assessed separately as it is influenced by the gene panel design, sequence data quality and a well matched reference data set. Jumble can also be applied to whole genome and other targeted sequencing such as exome, for which performance is not assessed in this study.

## Data Availability

All data relevant for the interpretation of our findings is provided in the main manuscript or the supplemental information. Dataset 1: Panel sequencing data, cfDNA and matched gDNA, from the ProBio trial (ClinicalTrials.gov identifier: NCT03903835, www.probiotrial.org). Any data providing information on individual outcomes or genotypes is considered to be a personal registry by the Swedish law (Personal Data Act), thereby prohibiting the submission of the sequencing files to a public repository. Access to the sequencing data data requires approval from the Swedish Ethical Review Authority and an agreement with the data protection- and legal unit at Karolinska Institutet. Dataset 2: Panel sequencing data of cfDNA from the MAGNITUDE trial (ClinicalTrials.gov identifier: NCT03748641). The data sharing policy is described in the publication, "Niraparib and Abiraterone Acetate for Metastatic Castration-Resistant Prostate Cancer", pmid 36952634. Dataset 3: Panel sequencing data and low-pass whole genome sequencing from a Danish retrospective cohort. See publication "Prognostic Value of Low-Pass Whole Genome Sequencing of Circulating Tumor DNA in Metastatic Castration-Resistant Prostate Cancer", pmid 36762756, for data access information.

