## Supplementary information for "Sensitive detection of copy number alterations in samples with low circulating tumor DNA fraction"

### Supplementary Figures

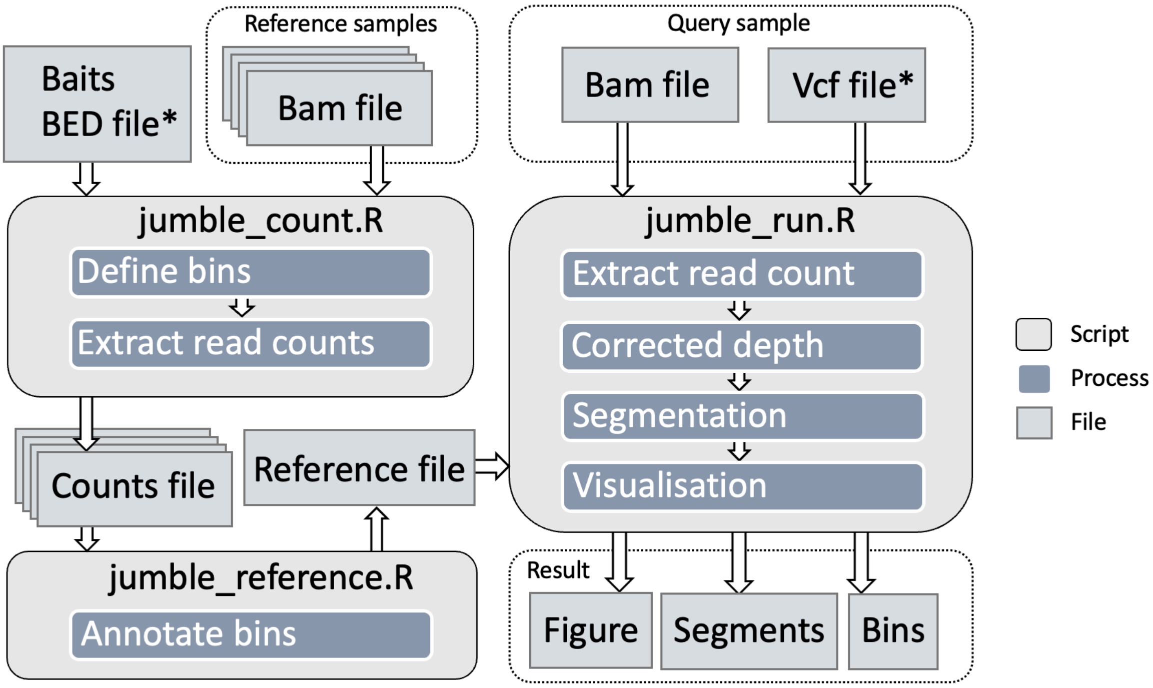


***Supplementary Figure 1. Schematic of the Jumble computational tool architecture.*** *Executable R scripts are run to process the reference and query samples. For a query sample, the main output consists of a figure file and two text files with tables of bins and segments. Asterisk indicates optional input.*

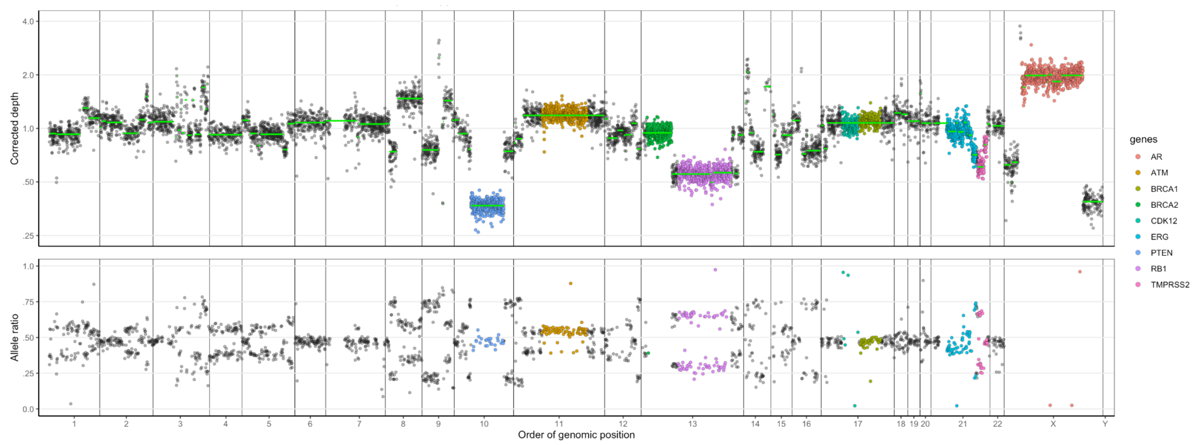


***Supplementary Figure 2. Homozygous deletion example.*** *Example of a copy number profile featuring homozygous deletion of PTEN (blue), identified as a deep, focal deletion featuring a SNP allele ratio near 0.5.* ***Upper panel:*** *Corrected sequencing depth serves as a measure of relative DNA abundance. Each dot is a genomic bin, positioned on the x axis in order along each chromosome of the reference genome. Corrected depth is centered around a point estimate such as mean or median, typically corresponding to 2 copies per cell in normal cells and about 2-4 (typically referred to as ploidy) for cancer cells. Increases in copy number relative to the ploidy appear at a corrected sequencing depth above one, and decreases in copy number appear below 1. Segmentation is shown in green.* ***Lower panel:*** *Alternative allele ratio of heterozygous SNPs by matchingtarget bin. A balanced copy number, with equal number of copies of each homologue, as for PTEN (blue), is indicated by SNP allele ratio near 0.5 (homozygously deleted, only normal DNA remains), and imbalanced copy number, such as for genes ATM (orange) and RB1 (purple) is indicated by a SNP allele ratio above or below 0.5, depending on whether the alternative allele is located on a retained/gained or lost/less amplified homologue.*

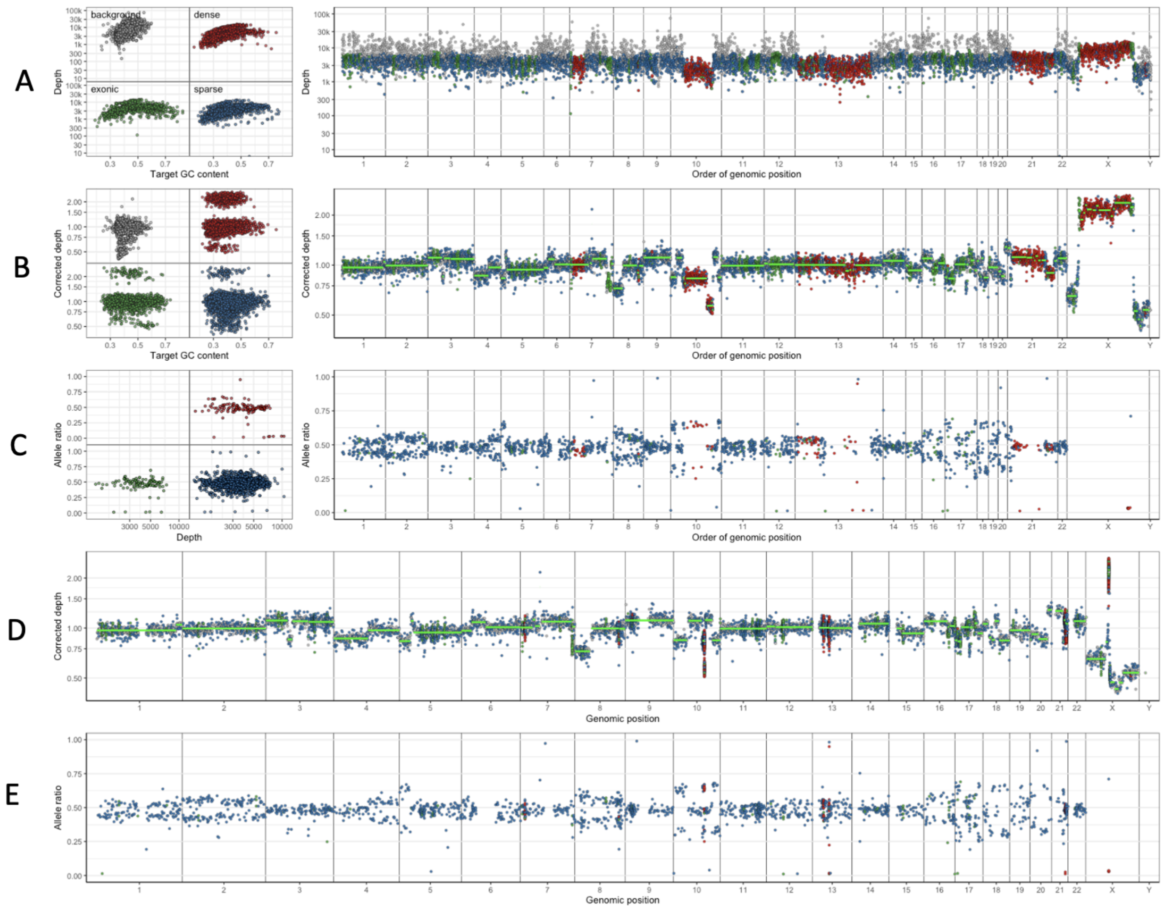


***Supplementary Figure 3. Overview of results from running Jumble with the ProBio panel.*** *At a ctDNA fraction near 0.4, a partial PTEN homozygous deletion (chromosome 10) and an AR amplification (chromosome X) are clearly visible.* ***A)*** *left: Sequence depth to GC content shows GC sequence bias over bins (background, densely targeted, exonic and other); right: Depth by bin and order of genomic position.* ***B)*** *left: corrected depth by GC content; right: corrected depth by order of genomic position.* ***C)*** *left: Heterozygous SNP allele ratio by sequence depth; right: SNP allele ratio by order of genomic position.* ***D)*** *Corrected depth by genomic position.* ***E)*** *SNP allele ratio by genomic position.*

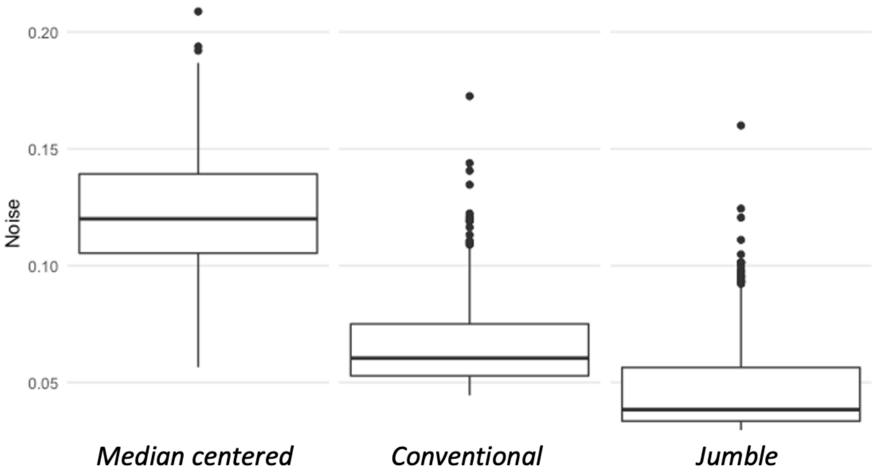


***Supplementary Figure 4. Noise levels.*** *Noise levels of raw (median-centered) sequence depth, conventional depth correction (subtraction of reference set median by target, and loess-based GC content correction) and Jumble depth correction (as described) in Dataset 2 (n=266). Noise was quantified as the median relative difference between adjacent target bins (higher divided by lower).*

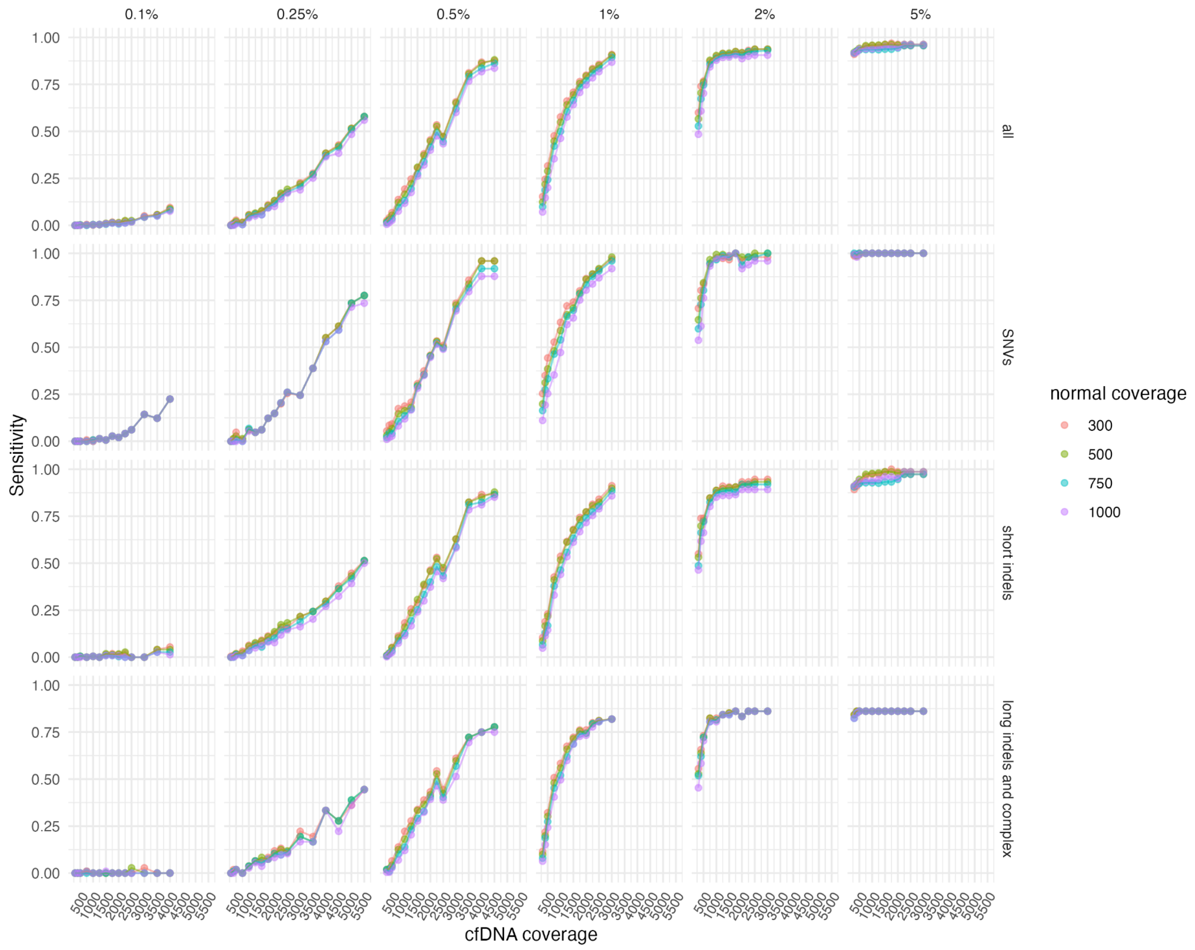


***Supplementary Figure 5. Sensitivity of somatic small variant calling versus mean target consensus coverage in ctDNA reference samples.*** *Somatic small variant calling was performed as described in Methods. The variants with high or moderate impact within the targeted regions were analyzed for sensitivity. Reference cfDNA samples (Twist Bioscience), with known variants in given variant allele fractions (VAF) were analyzed (0.1%, 0.25%, 0.5%, 1%, 2% and 5%) in 3-6 replicates, with the corresponding wildtype sample as a matched normal. BAM files were downsampled to desired coverage levels with samtools view. For highest tumor sample coverages, BAM files from replicates of the same VAF levels were pairwise merged and then downsampled. Columns show the different VAF levels. Rows show different variant types: all - all variants (N=159); SNVs - single nucleotide variants (N=40); short indels - insertions or deletions (indels) ≤5 base pairs (bp) (N=74); long indels and complex - indels ≥ 6 bp and complex variants (both reference allele and alternate allele is > 1 bp) (N=36). Sensitivity was calculated as the number of detected known variants of each type divided by the total number of known variants of the corresponding type within the targeted regions, and the mean was calculated over replicates.*

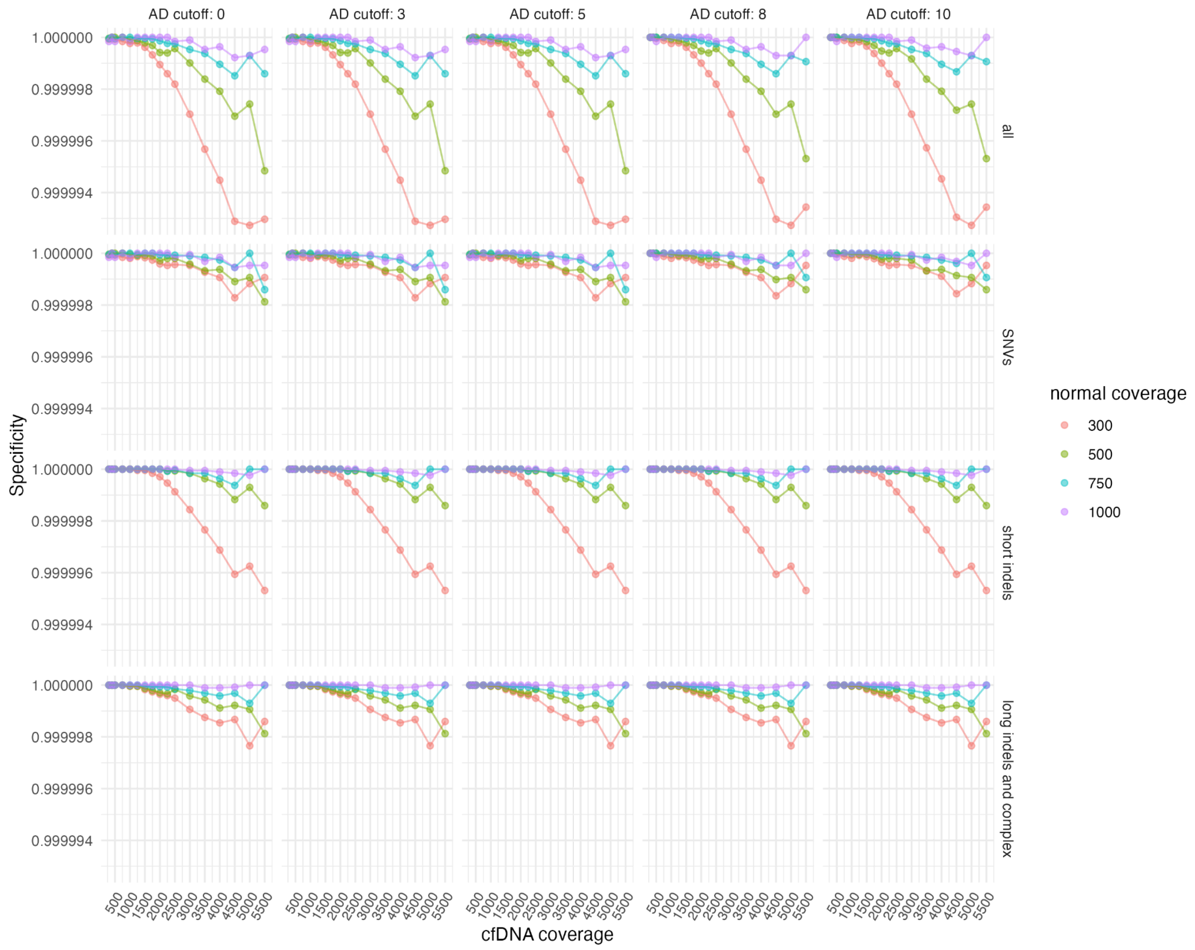


***Supplementary Figure 6. Specificity of somatic small variant calling versus mean target consensus coverage in ctDNA reference samples.*** *Somatic small variant calling was performed as described in Methods. The variants with high or moderate impact within the targeted regions were analyzed for specificity, with different tumor alternate allele depth (AD) cutoffs, shown in columns. Rows show different variant types: all - all variants; SNVs - single nucleotide variants; short indels - insertions or deletions (indels) ≤5 base pairs (bp); long indel and complex - indels ≥ 6 bp and complex variants (both reference allele and alternate allele is > 1 bp. 12 anonymous healthy donor (HD) blood samples were used, with cfDNA and matching white blood cell samples as normals. BAM files were downsampled to desired coverage levels with samtools view. For highest tumor sample coverages, BAM files from two HDs were pairwise merged, and the same for corresponding normal samples, and then downsampled. Specificity was calculated as 1 minus the total number of detected variants of each type divided by the total number of positions within the targeted regions, and the mean was calculated over samples.*

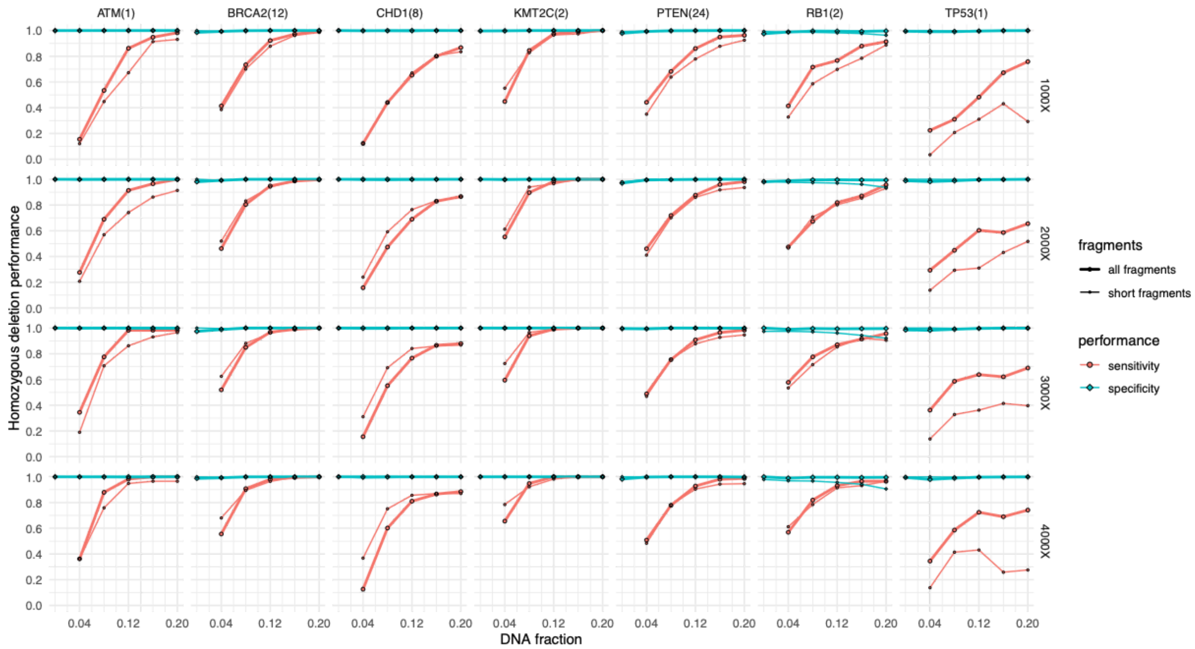


***Supplementary Figure 7. Sensitivity and specificity to detect homozygous deletions.*** *In silico dilution series of samples with high-confidence observation of homozygous deletion, each of them diluted with each of 58 ctDNA-negative samples (adding reads from another 1-2 ctDNA negative samples as required to avoid oversampling) to sequencing depths of 1000-4000. Sensitivity and specificity estimates are shown for the included genes, by ctDNA fraction, fragment length and sequencing depth. Filtering sequence reads by fragment length (<150 bp) did largely not improve performance. The number of ctDNA samples by gene with homozygous deletions, available for generating dilutions in the experiment, is shown in parenthesis, with the subsequent number of diluted samples therefore being 58 times that number.*

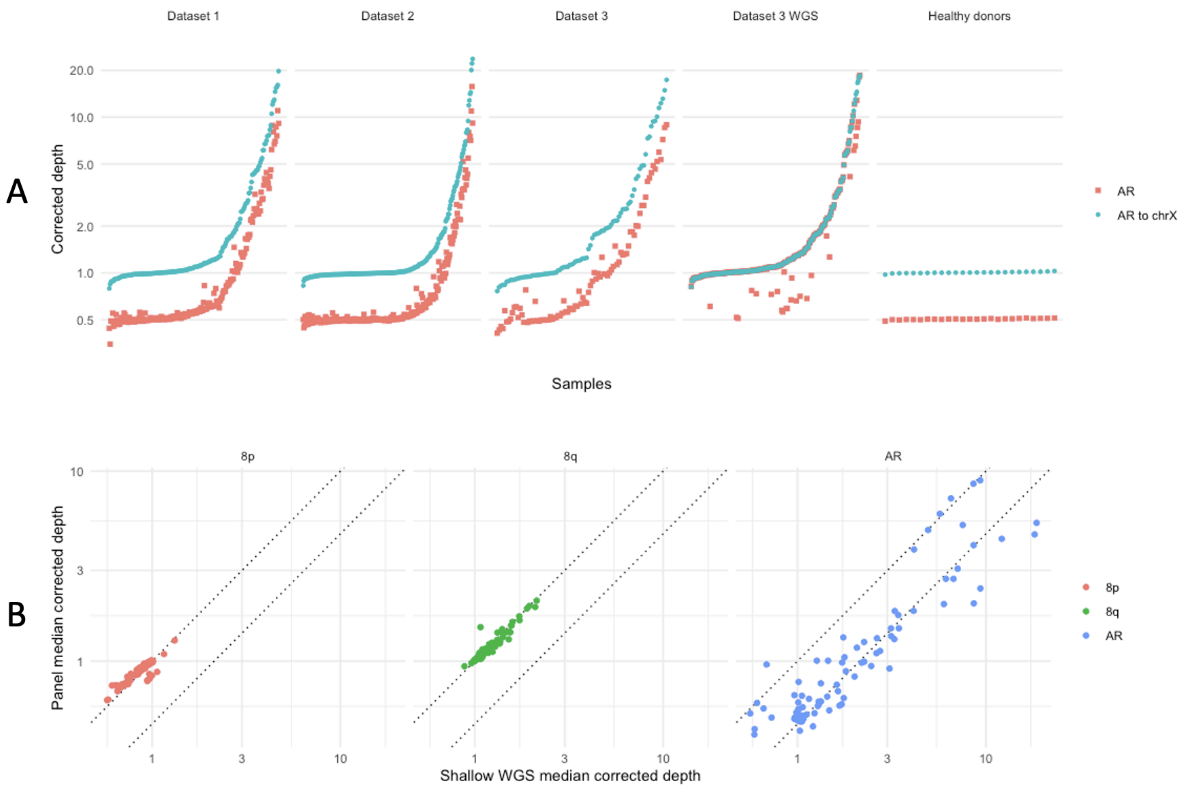


***Supplementary Figure 8. AR corrected depth distributions and comparison to low-pass whole genome sequencing data. A)*** *Corrected depth relative to sample median (red) and relative to chromosome X median (blue) for the AR gene in cfDNA from Dataset 1-3 (metastatic prostate cancer) and a set of 25 healthy donors. For Dataset 3, WGS corrected depth largely coincided with the corrected depth relative to chromosome X, as IchorCNA applied a haploid reference in most cases.* ***B)*** *Corrected depth (median per megabase) over Dataset 3, comparing shallow WGS and IchorCNA (x-axis) with the ProBio panel and Jumble (y-axis). Dashed line represents expected concordance (lower dashed line applies where IchorCNA assumes haploid reference for chromosome X). Concordance is good and no saturation of the signal is indicated for targeted sequencing relative to WGS. More noise is observed for AR as it represents a single megabase (one WGS data point) spanning the AR enhancer and AR.*

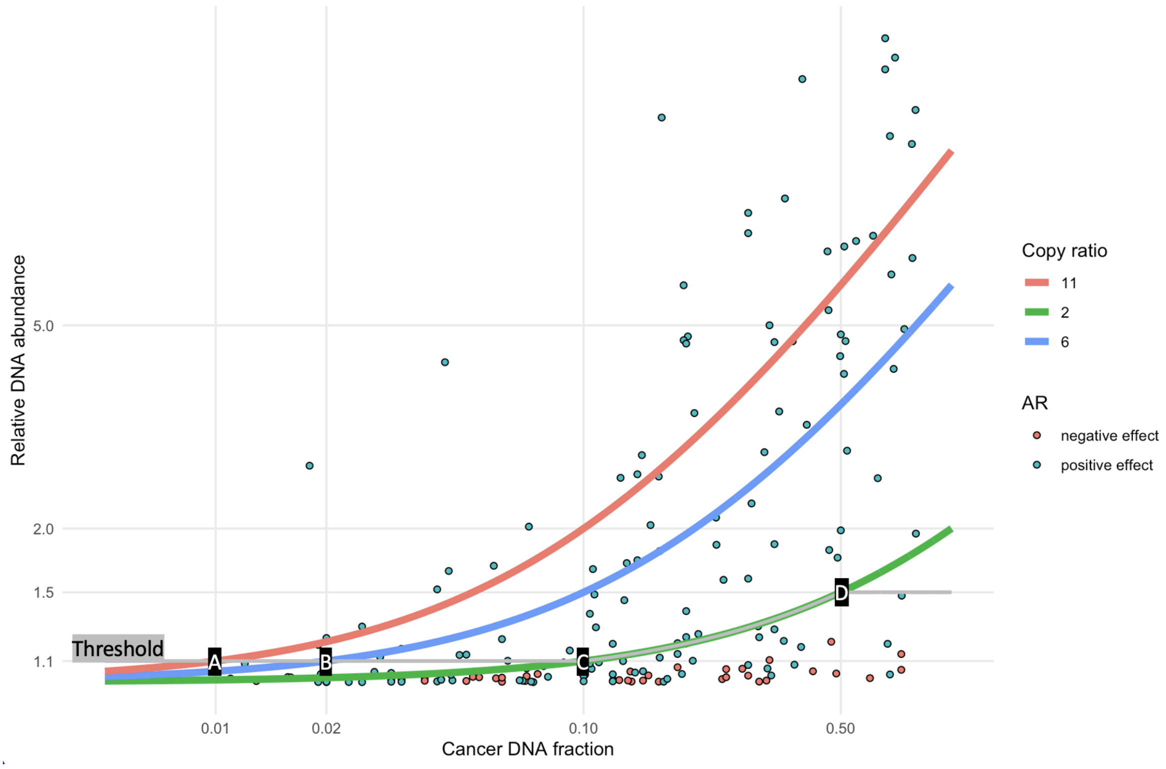


***Supplementary Figure 9. AR amplification sensitivity.*** *Overview of AR copy ratio observations in Dataset 1 (data points), expected signal by copy number (lines), and the set threshold for calling AR (and AR enhancer) amplifications (grey line).* ***A)*** *A threshold of 1.1 (grey line) corresponds to a limit of detection of 11 copies per cell (red line) at 0.01 cancer DNA fraction.* ***B)*** *At 6 copies per cell, the limit of detection is 0.02 cancer DNA fraction.* ***C)*** *At 0.1-0.5 cancer DNA fraction, the limit of detection is set to 2 copies per cell (green line), which is required to call amplification unless the relative DNA abundance is at least 1.5.* ***D)*** *A relative DNA abundance of ≥1.5 is enough to call amplification regardless of the resulting copy ratio. The observations are based on corrected AR depth (relative to 0.5, or to chromosome X median where that would yield a value closer to 1). Negative effects (below 1), were inverted and plotted as red points in the figure, representing noise/waviness that should not be called as amplification.*

Supplementary Tables

| Exons | Type | Gene-body sequencing | Focused CNV design | Intronic sequencing for fusions | Pharmacogenetic variants |
|---|---|---|---|---|---|
| AKT1 | HOTSPOT | AR | AR | TMPRSS2 | DPYD |
| BRAF | HOTSPOT | ATM | AR enhancer | ERG | HSD3B1 |
| CCND1 | HOTSPOT | BRCA1 | ATM | BRAF | HOXB13 |
| CDK4 | HOTSPOT | BRCA2 | BRCA2 | | COMT |
| CDK6 | HOTSPOT | CDK12 | CCND1 | | |
| CDKN1A | HOTSPOT | PTEN | CDK12 | | |
| CDKN2A | HOTSPOT | RB1 | CDKN2A | | |
| CTNNB1 | HOTSPOT | TP53 | CDKN2B | | |
| CUL3 | HOTSPOT | | CHD1 | | |
| DICER1 | HOTSPOT | | CHEK2 | | |
| FOXO1 | HOTSPOT | | FANCA | | |
| HRAS | HOTSPOT | | MYC | | |
| IDH1 | HOTSPOT | | NKX3-1 | | |
| KRAS | HOTSPOT | | PIK3CA | | |
| MED12 | HOTSPOT | | PIK3R1 | | |
| NRAS | HOTSPOT | | PTEN | | |
| PIK3CA | HOTSPOT | | RB1 | | |
| PIK3CB | HOTSPOT | | Intragenic region between TMPRSS2 and ERG | | |
| PIK3CD | HOTSPOT | | TP53 | | |
| PIK3R1 | HOTSPOT | | ZBTB16 | | |
| PIK3R2 | HOTSPOT | | | | |
| POLD1 | HOTSPOT | | | | |
| POLE | HOTSPOT | | | | |
| SF3B1 | HOTSPOT | | | | |
| SPOP | HOTSPOT | | | | |
| U2AF1 | HOTSPOT | | | | |
| XPO1 | HOTSPOT | | | | |
| APC | ALL_EXON | | | | |
| AR | ALL_EXON | | | | |
| ATM | ALL_EXON | | | | |
| BRCA1 | ALL_EXON | | | | |
| BRCA2 | ALL_EXON | | | | |
| CDK12 | ALL_EXON | | | | |
| DNMT3A | ALL_EXON | | | | |
| FOXA1 | ALL_EXON | | | | |
| KDM6A | ALL_EXON | | | | |
| KMT2A | ALL_EXON | | | | |
| KMT2C | ALL_EXON | | | | |
| KMT2D | ALL_EXON | | | | |
| PALB2 | ALL_EXON | | | | |
| PTEN | ALL_EXON | | | | |
| RB1 | ALL_EXON | | | | |
| TP53 | ALL_EXON | | | | |
| ARID1A | ALL_EXON | | | | |
| ARID2 | ALL_EXON | | | | |
| ATR | ALL_EXON | | | | |
| BARD1 | ALL_EXON | | | | |
| BRIP1 | ALL_EXON | | | | |
| CDH1 | ALL_EXON | | | | |
| CDKN1B | ALL_EXON | | | | |
| CHD1 | ALL_EXON | | | | |
| CHEK2 | ALL_EXON | | | | |
| FANCA | ALL_EXON | | | | |
| JAK1 | ALL_EXON | | | | |
| KEAP1 | ALL_EXON | | | | |
| MET | ALL_EXON | | | | |
| MGA | ALL_EXON | | | | |
| MLH1 | ALL_EXON | | | | |
| MLH3 | ALL_EXON | | | | |
| MRE11A | ALL_EXON | | | | |
| MSH2 | ALL_EXON | | | | |
| MSH3 | ALL_EXON | | | | |
| MSH6 | ALL_EXON | | | | |
| NBN | ALL_EXON | | | | |
| NCOR1 | ALL_EXON | | | | |
| NKX3-1 | ALL_EXON | | | | |
| PMS1 | ALL_EXON | | | | |
| PMS2 | ALL_EXON | | | | |
| RAD50 | ALL_EXON | | | | |
| RAD51 | ALL_EXON | | | | |
| RAD51B | ALL_EXON | | | | |
| RAD51C | ALL_EXON | | | | |
| RAD51D | ALL_EXON | | | | |
| RNF43 | ALL_EXON | | | | |
| SETD2 | ALL_EXON | | | | |
| SPEN | ALL_EXON | | | | |
| ZFHX3 | ALL_EXON | | | | |
| ZMYM3 | ALL_EXON | | | | |

| | | ProBio design |
|---|---|---|
| **Mutations** | | |
| | All coding exons | 51 genes |
| | Hotspots | 27 genes |
| **Pharmacogenetic variants** | | |
| | SNPs | 4 genes |
| **Copy-number alterations** | | |
| | Tiled SNP for genome-wide CNV | 3128 SNPs |
| | Directed analysis to increase sen | 20 genes |
| **Structural variation** | | |
| | Genefusions by intronic sequen | 3 genes |
| | Gene-body sequencing (e.g. BRC | 8 genes |
| **Microsatellite instability & Hypermutation** | | |
| | Microsatellites | 63 |
| | Hypermutation, entire design foc | Yes |
| | Associated genes | 6 |
| **DNA repair deficiency** | | |
| | Associated genes | 16 |
| **Total size (Mb)** | | |
| | | 1,6 |

*Supplementary Table S1. Content by gene of the ProBio panel.*

### Supplementary Methods

#### Depth correction algorithm

Read counts, or read pair (fragment) counts are generated, from aligned sequence reads, for all reference samples, and for each of a set of bins defined based on the sequence target specification (or the genome, for whole genome sequencing). Bin size is 200 bases for targeted regions of the genome, $10^6$ bases for untargeted regions, or $10^4$ default bases for WGS. The reference data set is stored in a reference data file, and is supplied to the analysis process each time a new query sample is analyzed. The reference samples need to be non-aberrant, i.e. not have copy number aberrations other than common polymorphisms. They should otherwise be similar to the query samples, e.g. processed with the same or similar tools and reagents, and sequence capture and panel design. Additionally, the reference samples should to a good extent span the biological and technical variability expected to occur in query samples analyzed.

For a query sample to be processed, read counts (referred to as sequencing depth) are extracted from aligned sequence reads based on the reference file definition, and corrected for systematic variability based on the reference sample set, as described in detail in below.

1. A reference set is built from a number of samples, at least one but preferably tens to hundreds.
2. From a BED (Browser Extensible Data) file defining targets or target baits, “target” bins (genomic regions) are defined to a length certain length, typically 200 bases, extending and/or merging each bait or target in both directions to a multiple of that length if needed, then splitting.
3. Background bins are also generated, with a length of about 1M bases, where they fit between target bins. These are processed similarly to target bins, as off-target reads over large genomic regions allow similar quantification of DNA abundance, at much lower resolution, where there are no target bins available.
4. In case of whole genome sequencing (WGS), no BED file of targets is relevant. Instead, one bin size is chosen and the entire reference genome is split into bins of that size. These are then processed as target bins.
5. A subset of bins are defined as *training*. These are located on autosomes and no more than a certain number, typically 10-50, of bins are used for each of all or a selection of genes. Thus all targeted autosomal genes should be present in the training set but the potential for genes to be overrepresented is limited. This limitation per gene is not applied with WGS.
6. For all reference samples, the number of reads or read pairs (DNA fragments) mapping to each bin is quantified from the BAM file of aligned sequence reads.
7. Read counts of zero are replaced with 1 to avoid log-scale issues.
8. For each reference file, raw log ratio (r*RLR*) is computed as:
$$rRLR = log2(\frac{readcount}{median(readcount_{training})})$$
9. For male reference samples, 1 is added to (non-pseudoautosomal) sex chromosome *RLR* (doubling the sequence depth). For female samples Y chromosome r*RLR* are drawn from a normal distribution resembling autosomal r*RLR*. This is to allow subsequent application of the reference set to give results relative to “normal diploid reference” for all chromosomes.
10. Bins with predominantly zero or very low, high or variable r*RLR* are removed from the reference set and not used in further analysis.

11. For the query sample to be analyzed for copy number aberrations, the number of reads or read pairs (DNA fragments) mapping to each bin is quantified from the BAM file of aligned sequence reads.
12. Read counts of zero are replaced with 1.
13. Query sample raw log ratio (*RLR*) is computed as:

$$RLR = log2(\frac{readcount}{median(readcount_{training})})$$

14. Principal component analysis (PCA) is applied to r*RLR* to compute scores for each principal component (PC) and bin (i.e. not per sample). The PCs represent a set of *latent features* (capturing effects such as GC content) that affect sequence coverage, and each score represents the relative amount by which this feature influences the r*RLR* of that bin in the reference samples.
15. Outlier bins, based on r*RLR* PCA scores, are removed from further analysis. An example of an outlier bin would be where local GC content, mappability, or some other feature affecting sequence coverage, is very high or low. Bins with a PCA score above or below a certain number of standard deviations (typically 4), for any of the PCs, are considered outliers and removed.
16. PCA is reapplied to the retained set of r*RLR* to improve (reduce the effect of potential outliers) the PCA and the latent features.
17. For the first or several first few PCs, an estimate of query sample *RLR* systematic variability *error* is estimated from a regression model *m*. To reduce potential confounding of error with signal, only the *training RLR* of the query sample is used to build the model:

$$m = RLR_{training} \sim scores_{training,PC1,..}$$

$$error = predict(m,\ scores_{PC1,..})$$

18. The error is then removed from the *RLR* to create a corrected log ratio (*LR*):

$$LR = RLR - error$$

19. The above two steps are repeated, with a new model and correction performed iteratively using the already corrected *LR* and some or all of the remaining PCs:

$$m = LR_{training} \sim scores_{training,PCi,..}$$

$$error = predict(m,\ scores_{PCi,..})$$

$$LR = LR - error$$

20. To make sure GC content bias has been adequately captured and corrected for in the query sample, an explicit correction step for GC content can be applied. Here, a LOESS model is used. Again, only *training* bins are used when training the model:

$$m = LR_{training} \sim GCcontent_{training}$$

$$error = predict(m,\ GCcontent)$$

$$LR = LR - error$$

21. For targeted sequencing, *LR* of background (not to be confused with *training*) bins is computed separately, using the same procedure.
22. Corrected depth for all bins is $2^{LR}$ but typically displayed on the log scale. The *LR* is preferred in some downstream steps such as segmentation, while the corrected depth is more intuitive to interpret.

#### Dilution experiment

A dilution series of ctDNA samples representing tumor DNA content fractions of 0, 0.04, 0.08, 0.12, 0.16 and 0.20 was generated from a subset of Dataset 2, using all ctDNA samples carrying an obvious homozygous deletion (n=44) affecting genes *ATM* (n=1), *BRCA2* (n=12), *CHD1* (n=8), *KMT2C* (n=2), *PTEN* (n=24), *RB1* (n=2) and *TP53* (n=1), and all samples considered ctDNA negative based on comprehensive genomic analysis and a median sequence coverage above 1500 (n=58). Tumor DNA fractions were estimated assuming a simple linear relationship between the tumor DNA fraction and the effect on corrected depth observed for homozygous deletion ($fraction = 1 - corrected\ depth$). Sequence coverage was calculated for each sample from the median sequence depth (DP) over autosomal SNPs. For each combination of tumor and normal sample, a dilution was created at the aforementioned ctDNA fractions and at sequence depths (based on median SNP depth) of 1000, 2000, 3000 and 4000X. Fragment counts per bin, using Jumble, were subsampled from each source sample pair using a binomial randomization (R, rbinom) with probabilities calculated from the tumor DNA fraction of the tumor sample and the aforementioned sequence depth (from SNPs) of both source samples. Fragment counts for fragments shorter than 150 bases were also generated. Where the normal DNA would need to be oversampled due to insufficient sequence depth, to achieve the specified sequence depth, the full source fragment count was used, and more were sampled from a second normal sample, and similarly using third normal if necessary. Where the tumor sample would need to be oversampled due to insufficient combination of sequence depth and tumor DNA fraction, oversampling was performed by including the full source fragment count, and sampling additional counts from the same sample. Each diluted sample was saved as a Jumble “count” file and further processed with Jumble using the reference file generated from Dataset 1, as if it had been parsed from a BAM file. Results from Jumble were segmented using CBS (R, PSCBS package, alpha=0.01, undo=0) and segments were considered putative focal deletion calls, for each included gene, if the following criteria were met: 1) one or more segments were generated for the relevant gene; 2) the number of bins included in the segment(s) were at least 20, or 90% of that of the smallest real deletion of that gene included in the dilution series, if that was a lower number; and 3) the signal effect of the segment(s) exceeded ⅔ of the expected, given the tumor DNA content and a homozygous deletion. For each diluted sample, the result per gene was summarized as true positive, true negative, false positive or false negative based on whether the tumor sample carried a real homozygous deletion of that gene. Sensitivity and specificity were summarized per gene, tumor fraction and sequence coverage. Sensitivity was not defined where tumor DNA fraction was zero. Thus, for each tumor DNA fraction, gene and coverage, sensitivity was based on samples with a real homozygous deletion of that gene, and specificity was based on the samples without. The same analysis was carried out using only the shorter fragments, for which the same fragment length filter was applied to the reference samples.
